# Feasibility of PI-RADS Compliant Prostate MRI at 0.55T

**DOI:** 10.64898/2026.05.05.26352358

**Authors:** Arthur P C Spencer, Emanuele Avola, Gorun Ilanjian, Jade Matthey, Jean-Baptiste Ledoux, Clarisse Dromain, Naïk Vietti-Violi, Ileana Jelescu

## Abstract

**Purpose:** To demonstrate the feasibility of prostate MRI at low-field and provide an optimised low-field prostate MRI protocol.

**Methods:** In 15 healthy male volunteers (mean age 62 years, 95% CI 57–68) and 22 patients with suspected prostate cancer (mean age 68 years, 95% CI 64–71), we acquired bi-parametric prostate MRI at 0.55T including tri-planar T2-weighted imaging (WI) and axial diffusion weighted imaging (DWI). Parameters adhered to PI-RADS guidelines and deep learning reconstruction (DLR) was used to enhance image quality. In healthy volunteers, we optimised the protocol based on evaluation of SNR, CNR and qualitative image scores. In patients, we evaluated qualitative image scores and lesion detection performance using 3T as the reference.

**Results:** The acquisition time (minutes:seconds) was 4:02 for axial T2-WI and 6:32 for DWI. In healthy controls, DLR substantially improved T2-WI quality (p<0.05). The optimised DWI protocol had b-values of 50 and 800 s mm^-2^, with a calculated b-value image at 1500 s mm^-2^. Twenty-one patients (95%), and all controls, had at least acceptable diagnostic quality (PI-QUAL score≥2/3) at low-field MRI. With respect to the 3T reference, two readers detected lesions at low-field with sensitivity of 79% and 75% at the lesion level. At the patient level, the sensitivity was 91% for both readers. Low-field patient-level lesion detection did not significantly differ from 3T (p=0.5).

**Conclusion:** Prostate MRI is feasible at low-field, providing clinically acceptable acquisition times for diagnostic quality images with comparable patient-level lesion detection performance to 3T.

## Introduction

Prostate cancer (PCa) is a leading cause of cancer-related mortality in males [1]. The recent adoption of magnetic resonance imaging (MRI) has transformed the clinical management of PCa, facilitating MRI-targeted biopsies which improve detection of clinically significant PCa while reducing overdiagnosis of indolent lesions [2].

The Prostate Imaging Reporting and Data System (PI-RADS) recommends the use of 3T MRI for prostate imaging due to its superior signal-to-noise ratio (SNR) compared to lower fields [3]. 1.5T examinations remain acceptable in specific cases, such as the presence of MRI-conditional implants or metallic implants (e.g. hip prostheses) [3]. The use of MRI below 1T is not yet recommended due to the lack of clinical validation [3]. Nevertheless, low-field MRI scanners offer notable economic and technical advantages. Specifically, lower acquisition cost and decreased maintenance requirements [4], as well as reduced specific absorption rate, diminished susceptibility artifacts, and improved patient comfort due to larger bore diameters [5], make them a viable and affordable solution. However, the main drawback remains the lower SNR compared to higher field strengths [5]. To address the need for standardised image quality assessment, the Prostate Imaging Quality (PI-QUAL) score evaluates prostate MRI quality using objective criteria [6], based on T2-weighted imaging (T2-WI), diffusion-weighted imaging (DWI), and apparent diffusion coefficient (ADC) map [7].

The potential of low-field MRI in prostate imaging is increasingly recognised [8] but remains mostly unexplored. A recent study showed similar volumetric assessment based on T2-WI and similar ADC map values between 3T and 0.55T prostate exams [9]. Additionally, MRI-guided prostate biopsy has been performed at 0.55T, but relied on 3T examinations for lesion localisation [10]. Recent studies have assessed the potential of 0.55T MRI for abdominal imaging [5, 11], reporting a diagnostic quality comparable to higher field strengths [4, 12].

Additionally, deep learning-based image reconstruction (DLR) techniques have shown potential to enhance SNR in low-field MRI [11, 13] and in prostate MRI at 3T [14–18], but their utility for prostate MRI at low-field has not been evaluated. To date, no studies have evaluated SNR and contrast-to-noise (CNR), reported a qualitative assessment using the PI-QUAL score, nor evaluated lesion detection performance at 0.55T.

In this study, we developed and optimised a clinically feasible protocol for prostate MRI at 0.55T. We assessed qualitative and quantitative measures of image quality, including SNR and CNR on T2-WI and DWI and their derivatives, and PI-QUAL score. We then evaluated the lesion detection performance of the proposed low-field protocol in comparison with 3T.

## Methods

### Study design

This prospective study was approved by the local ethics committee (CER-VD no. 2024-D0049). This study was performed in two phases. First, healthy control data was used for quantitative and qualitative image quality assessment and protocol optimisation. Then, the optimised MRI protocol was acquired in suspected PCa patients to evaluate lesion detection performance.

Fifteen healthy male volunteers (>50 years old) with no clinical suspicion of PCa, and 22 suspected PCa patients (adult male), underwent low-field prostate MRI between September 2024 and June 2026. Written informed consent was obtained from all participants prior to imaging. Suspected PCa patients were selected following identification of one or more PI-RADS 4 or 5 lesions at 3T clinical prostate MRI. Patients with a history of PCa were excluded.

### MRI Protocol

Low-field MRI examinations were conducted using a 0.55T system (MAGNETOM Free.Max, Siemens Healthineers, Forchheim, Germany) equipped with a medium 12-channel flexible surface coil. The position on the table was feet first supine, and a wide belt was installed on the pelvis to reduce artifacts related to breathing. No intravenous contrast or antispasmodic agent was administered, and scans were conducted without prior enema. We acquired a PI-RADS bi-parametric MRI protocol (tri-planar T2-WI and axial DWI) in addition to an axial non-contrast T1-WI sequence. Acquisition parameters (Table 1) adhered to the recommendations described in the PI-RADS version 2.1 guidelines [19].

**Table 1:** Low-field MRI acquisition parameters.

|  | **T2-WI**<br>(axial / sagittal /<br>coronal) | **DWI** [50, 800] | **DWI** [50, 1000] | **T1-WI** |
| --- | --- | --- | --- | --- |
| TR (ms) | 7580 / 6970 / 7150 | 4000 | 4100 | 773 |
| TE (ms) | 114 | 100 | 102 | 10 |
| Resolution (mm) | 0.4 x 0.4 | 2 x 2 (acquired)<br>1 x 1 (super-resolution<br>reconstruction) |  | 0.6 x 0.6 |
| Slice thickness (mm) | 3.0 | 3.5 |  | 4.0<br>(+20% slice gap) |
| Matrix | 512 x 512 | 86 x 86 (acquisition matrix)<br>172 x 172 (super-resolution<br>reconstruction) |  | 384 x 608 |
| Field of view (mm) | 200 x 200 | 177 x 177 |  | 240 x 380 |
| Number of slices | 26 / 25 / 25 | 18 |  | 50 |
| Acceleration | GRAPPA 4 | Partial Fourier 6/8 |  | GRAPPA 3 |
| Turbo factor | 25 | n/a |  | 3 |
| b-values (s mm<sup>-2</sup>) | n/a | 50, 800 | 50, 1000 | n/a |
| Number of signal<br>averages | 4 | 4, 20 | 4, 20 | 1 |
| Acquisition time<br>(minutes : seconds) | 4:02 / 3:43 / 3:48 | 6:32 | 6:42 | 2:34 |

Axial T1-WI were acquired with a turbo spin echo sequence. Tri-planar T2-WI were acquired with a turbo spin-echo sequence, and DLR was performed using ‘medium’ Deep Resolve, as recommended by the vendor (Siemens Healthineers). For the control subjects, reconstructions without DLR were stored for analysis of SNR improvements offered by DLR.

Axial DWI data were acquired with an echo-planar imaging research sequence. DLR was applied using the vendor’s implementation of Deep Resolve (Siemens Healthineers) [11, 20]. This comprised denoising with hyperparameters optimised for enhanced denoising to mitigate the low SNR encountered on Free.Max, and super-resolution reconstruction using a dedicated approach for partial Fourier acquisition. Retrospective reconstructions without DLR were not available for DWI data. The acquired voxel size was 2 x 2 x 3.5 mm^3^, giving voxels of 1 x 1 x 3.5 mm^3^ after super-resolution reconstruction. In a subset of healthy control subjects (n = 11), we acquired two DWI datasets: one at b-values of 50 and 1000 s mm^-2^ (DWI_1000_) consistent with a 3T clinical protocol, and another at b-values of 50 and 800 s mm^-2^ (DWI_800_), each with six directions per b-value. The b-value of 800 s mm^-2^ was eventually selected to improve SNR compared to 1000 s mm^-2^, in order to compensate for the lower SNR of low-field MRI. For the remaining control subjects (n = 4) and patients (n = 22), only the DWI_800_ protocol was acquired. For each acquisition, trace-weighted images at each b-value were derived, in addition to ADC maps, and calculated b-value images for 1400–2000 s mm^-2^ in steps of 100 s mm^-2^ in order to also evaluate the optimal calculated b-value for screening.

Suspected PCa patients also underwent prostate MRI with a clinical protocol on a 3T system (MAGNETOM VidaFit, Siemens Healthineers) following the PI-RADS version 2.1 guidelines [19]. Tri-planar T2-WI were acquired with a turbo spin-echo sequence using DLR (Deep Resolve Boost, Siemens Healthineers). Axial DWI data were acquired with an echo-planar imaging sequence (ZoomIt, Siemens Healthineers), using b-values of 50 and 1000 s mm^-2^ with four directions per b-value. Complete acquisition parameters at 3T are given in Supplementary Table 1.

### Quantitative Image Assessment

Quantitative image assessment was performed on healthy control data to evaluate the improvements offered by DLR, and to enable optimisation of the DWI protocol. Whole prostate, transitional zone (TZ) and peripheral zone (PZ) 3D segmentations were performed automatically using Mint software on T2-WI. They were then reviewed by a radiologist with 7 years of expertise in prostate imaging (NVV) and corrected if needed. For each subject, the ADC map was nonlinearly registered to the axial T2-WI sequence using Elastix [21, 22]. The inverse of the resulting transformation was used to align the prostate segmentation with the DWI images.

Apparent SNR was calculated within the whole prostate, TZ and PZ using [23]:

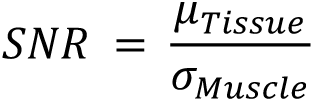

where *μ_Tissue_* is the mean image intensity in the tissue (whole prostate, TZ, or PZ), and *σ_Muscle_* is the standard deviation of the image intensity in a region of interest in the internal obturator muscle. To quantify the contrast between the PZ and TZ, the apparent CNR was calculated using [23]:

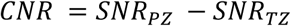

### Qualitative Image Assessment

Qualitative image assessment was performed on healthy control data to evaluate the improvements offered by DLR and to enable protocol optimisation, and on PCa patient data to enable comparison between low-field and 3T. Quality assessment was conducted by two board-certified radiologists with 7 years (NVV) and 5 years (EA) of experience in prostate MRI, using the PI-QUAL version 2 criteria for bi-parametric prostate MRI [7]. Both readers were blinded to each other’s evaluations. In case of disagreement between readers, a consensus score was established following a joint image review.

In healthy controls, qualitative analysis was performed on T2-WI with and without DLR for comparison, and on DWI_800_ and DWI_1000_ data for comparison. For each DWI sequence, scores were assigned based on the ADC map and the calculated b-value image with highest SNR in quantitative analysis (b = 1500 s mm^-2^ for DWI_800_ and b = 1400 s mm^-2^ for DWI_1000_). A final PI-QUAL score was assigned based on the scores from the optimised protocol (consisting of T2-WI with DLR and DWI_800_).

In patients, qualitative analysis was performed at low-field on T2-WI with DLR and DWI_800_ using the calculated b-value image at b = 1500 s mm^-2^, and at 3T on T2-WI and DWI using the calculated b-value image at b = 2000 s mm^-2^.

### Lesion Detection

At low-field, lesion detection was conducted by two board-certified radiologists with 7 years (NVV) and 5 years (EA) of experience in prostate MRI. Data from patients and healthy controls was pooled to provide a dataset with both true positives and true negatives. Readers were blinded to case-control status and to each other’s evaluations. At 3T, the lesion evaluation was performed by a consensus of both readers reporting PI-QUAL score and PI-RADS 4 or 5 lesions. Up to three lesions were detected per subject.

### Statistical Analysis

To assess the improvement offered by DLR on T2-WI, one-tailed Wilcoxon signed-rank tests were used to assess increases in SNR, CNR and qualitative image scores with DLR vs without DLR. To evaluate the optimal DWI acquisition, SNR and CNR values were compared between trace-weighted images acquired at b = 800 and 1000 s mm^-2^, and ADC values calculated from each acquisition were compared using two-tailed Mann-Whitney U tests. Qualitative scores were also compared between DWI_800_ and DWI_1000_ using two-tailed Mann-Whitney U tests. In patients, qualitative scores were compared between low-field and 3T using Mann-Whitney U tests. Bonferroni-corrected p-values < 0.05 were considered significant. To assess interrater agreement in qualitative image scores, squared weighted Cohen’s kappa was measured separately for patients and controls. In controls, Cohen’s kappa was measured for scores pooled across DWI_800_ (n = 15), DWI_1000_ (n = 11), T2-WI with DLR (n = 15) and T2-WI without DLR (n = 10) to provide sufficient statistical power (n = 51). In patients, Cohen’s kappa was measured for scores pooled across DWI (n = 22) and T2-WI (n = 22).

Lesion detection performance was evaluated at the lesion level (quantifying the number of individual lesions correctly identified) and the patient level (quantifying the number of subjects identified as having at least one lesion), using the 3T evaluation as the reference. We measured the lesion-level sensitivity for each rater, and the patient-level sensitivity and specificity for each rater. Patient-level lesion detection was statistically compared between low-field and 3T using an exact McNemar’s test. We measured Cohen’s kappa for the interrater agreement of patient-level lesion detection across patients and controls (total n = 37).

## Results

Healthy male volunteers (n = 15, mean age 62 years, 95% CI 57–68) and patients with suspected PCa (n = 22, mean age 68 years, 95% CI 64–71) were evaluated. The scan duration (minutes:seconds) for the PI-RADS bi-parametric protocol (Figure 1) was 4:02 for axial T2-WI and 6:32 for DWI. All scan durations are shown in Table 1.

**Figure 1:**
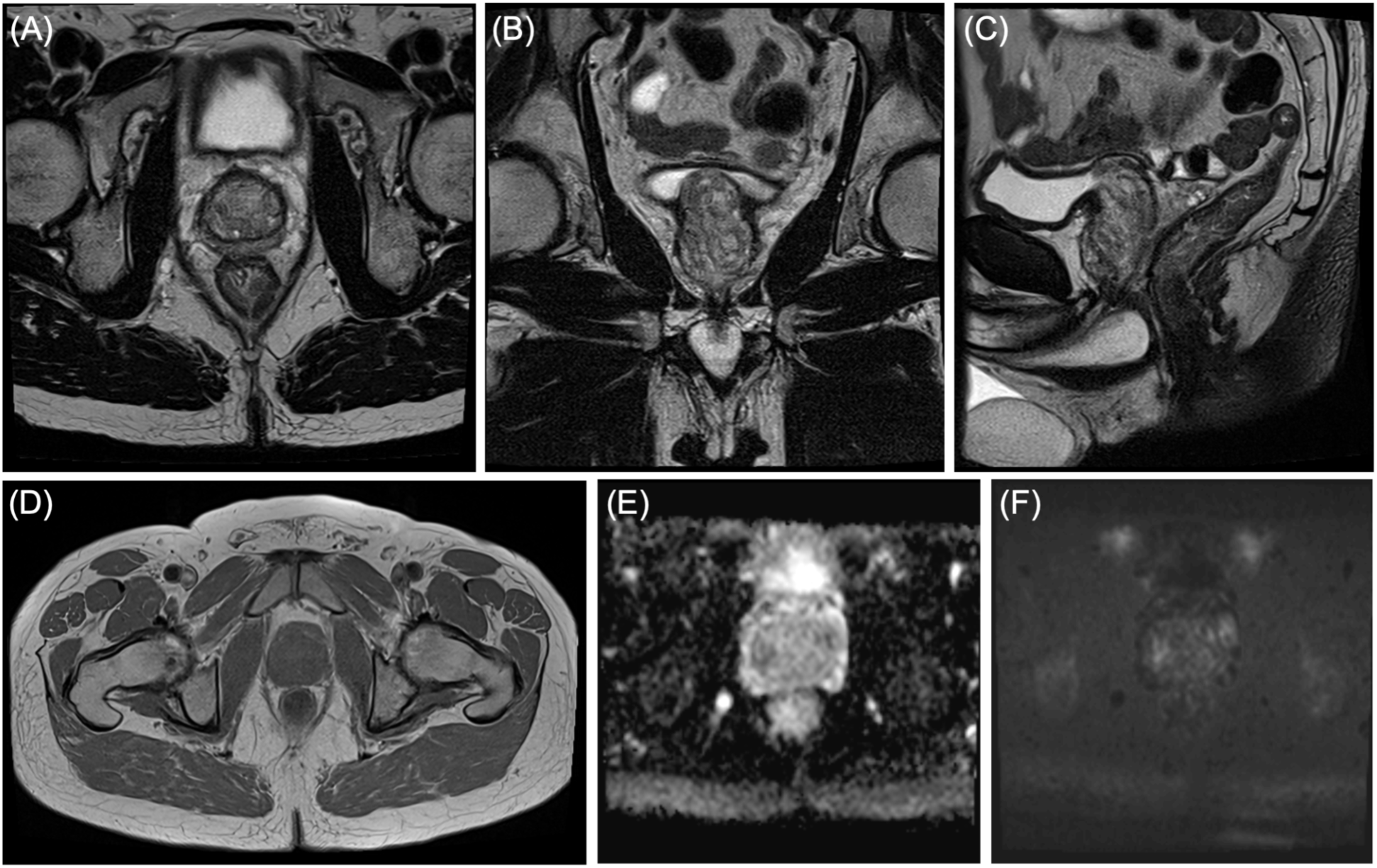
Representative example of the optimised MRI protocol in a healthy control, including axial T2-WI (A), coronal T2-WI (B), sagittal T2-WI (C), axial T1-WI (D), ADC (E), and calculated b-value image at b = 1500 s mm^-2^ (F). DWI derivatives were calculated from data acquired with b-values of 50 and 800 s mm^-2^.

### Quantitative Image Assessment

In data from healthy controls, axial T2-WI with DLR had a median (IQR) SNR of 10.86 (9.82– 11.93) in the whole prostate, 8.72 (7.27–9.67) in the TZ, and 12.84 (12.00–15.87) in the PZ (Figure 2E). Axial T2-WI without DLR had a median (IQR) SNR of 6.12 (5.59–6.63) in the whole prostate, 5.13 (4.68–5.68) in the TZ, and 7.45 (6.35–8.86) in the PZ (Figure 2E). CNR between the PZ and TZ was 5.84 (4.06–6.82) (Figure 2F). Where raw data were available for comparison (n = 10), DLR substantially improved image quality (p < 0.05; Figure 2).

**Figure 2:**
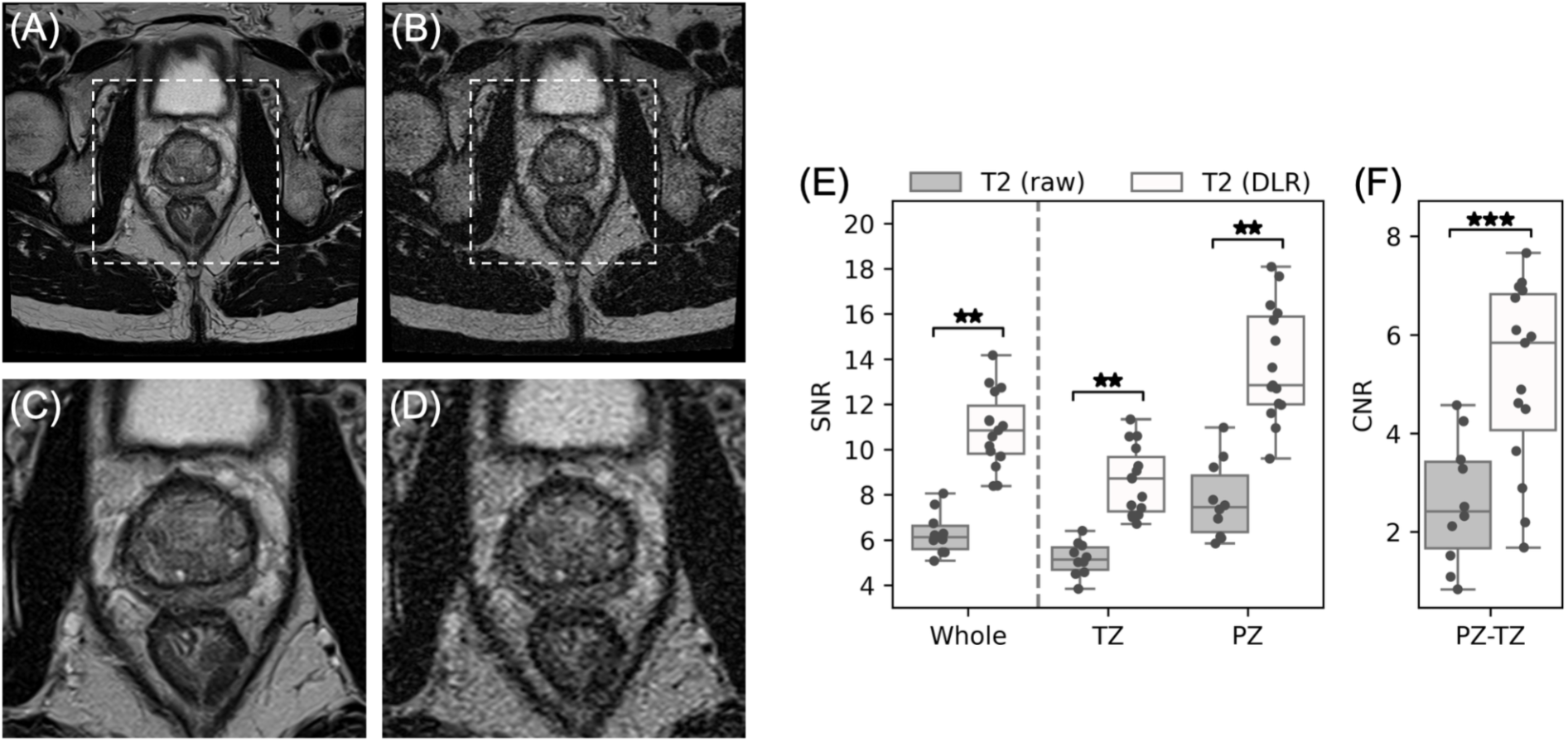
Representative example of axial T2-WI with DLR (A) and without DLR (B). The dashed boxes indicate the zoomed-in areas shown with DLR (C), and without DLR (D). Measurements of SNR (E) and CNR (F) from axial T2-WI, are shown for the whole prostate, TZ and PZ, with and without DLR. *p < 0.05; **p < 0.01; ***p < 0.001.

SNR was significantly higher in trace-weighted images at b = 800 s mm^-2^ compared to 1000 s mm^-2^ (p < 0.05; Supplementary Figure 1 and 2; Figure 3). There was no significant difference between ADC values calculated from b-values of 50 and 800 s mm^-2^ compared to those calculated from b-values of 50 and 1000 s mm^-2^. In calculated b-value images from DWI_800_, SNR was highest at b = 1500 s mm^-2^ (median 12.89, IQR 10.56–13.50; Figure 3G), and ADC offered good contrast between the PZ and TZ, with median ADC values of 1.70 and 1.44 x10^-3^ mm^2^ s^-1^ respectively (Figure 3H). In calculated b-value images from DWI_1000_, SNR was highest at b = 1400 s mm^-2^ (median 11.75, IQR 10.88–14.33; Supplementary Figure 2C).

**Figure 3:**
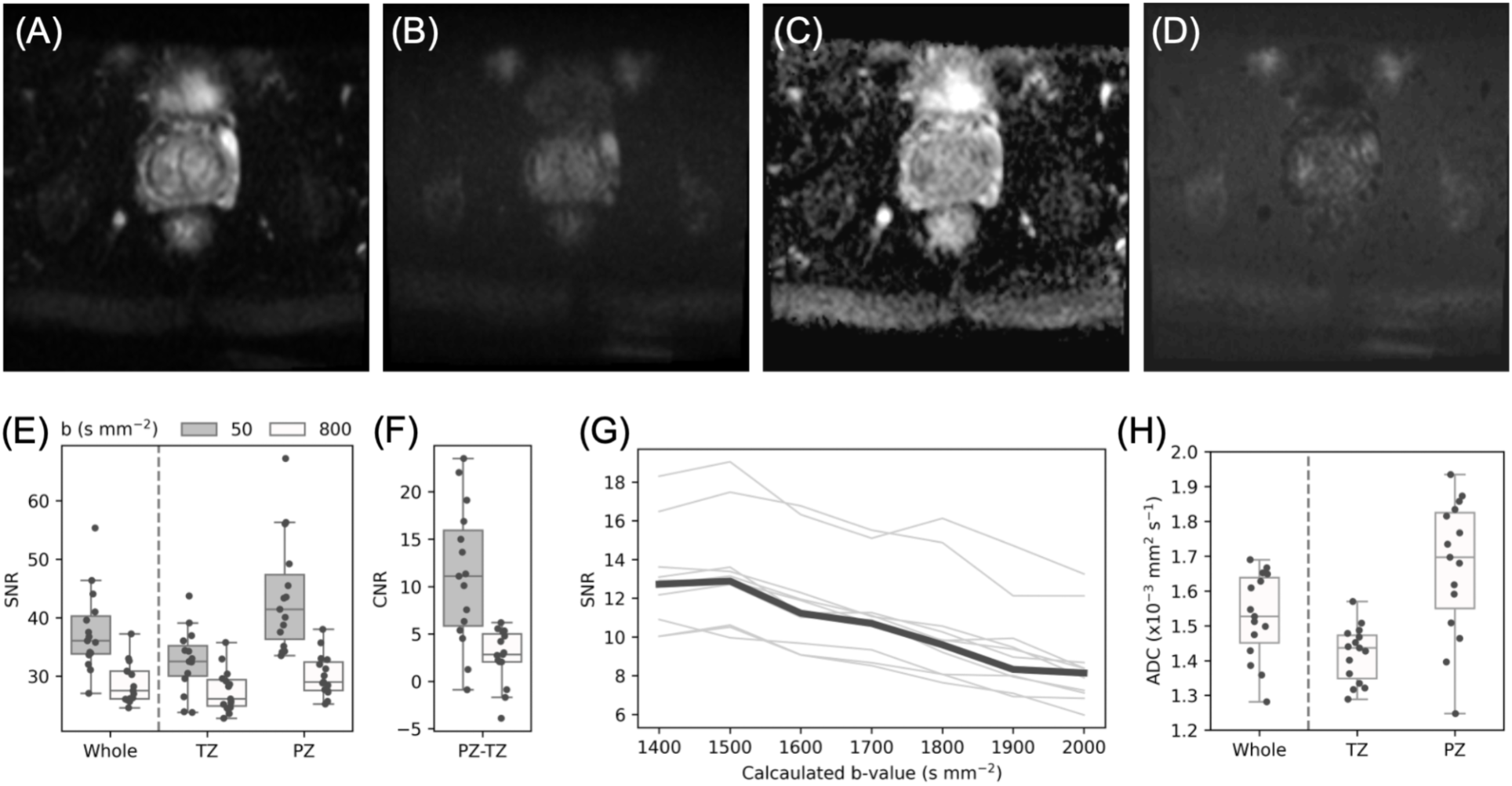
DWI data acquired with b-values of 50 and 800 s mm^-2^. Representative examples of DWI data are shown for b = 50 s mm^-2^ (A), b = 800 s mm^-2^ (B), ADC (C), and the calculated b-value image at b = 1500 s mm^-2^ (D). SNR (E) and CNR (F) are shown for each acquired b-value for the whole prostate, TZ and PZ. SNR in the whole prostate is shown for calculated b-values in the range 1400–2000 s mm^-2^ (G), showing each subject as a thin grey line, with the median shown as a thick black line. ADC values are shown for the whole prostate, TZ and PZ (H).

### Qualitative Image Assessment

Image quality scores for healthy controls at low-field are shown for each rater in Supplementary Table 2, with consensus scores shown in Supplementary Table 3. For T2-WI, image quality was higher with DLR than without (p = 0.002; Supplementary Table 3), improving scores from 3 to 4 in all ten participants. For DWI, there was a non-significant trend towards higher image quality scores for DWI_800_ compared to DWI_1000_ (p = 0.055; Supplementary Table 3). Both qualitative and quantitative results indicated the optimal bi-parametric protocol consisted of T2-WI with DLR and DWI_800_. Consensus scores, including overall PI-QUAL scores, are shown for these data in Table 2. Ten participants had an overall PI-QUAL score of 3/3 (indicating optimal diagnostic quality) and five participants had a score of 2/3 (indicating acceptable diagnostic quality). These five participants had a DWI score of 3/4 due to artifacts related to rectal gas (4 cases) or the presence of hip prostheses (1 case).

**Table 2:** Qualitative image scores in healthy controls for the optimised bi-parametric protocol comprising T2-WI with DLR and DWI with b-values of 50 and 800 s mm^-2^.

|  | Score | n (%) |
| --- | --- | --- |
| T2-WI | 0 | 0 (0) |
|  | 1 | 0 (0) |
|  | 2 | 0 (0) |
|  | 3 | 1 (7) |
|  | 4 | 14 (93) |
| DWI | 0 | 0 (0) |
|  | 1 | 0 (0) |
|  | 2 | 0 (0) |
|  | 3 | 5 (33) |
|  | 4 | 10 (67) |
| PI-QUAL | 1 | 0 (0) |
|  | 2 | 5 (33) |
|  | 3 | 10 (67) |

Image quality scores in suspected PCa patients at low-field are shown for each rater in Supplementary Table 4, and consensus scores in comparison with 3T are shown in Table 3. All patients had maximum image quality scores at 3T (Table 3). Compared to these, image quality scores in patients at low-field were not significantly different for T2-WI (p = 0.4859), but were significantly lower for DWI (p = 0.0140) with one patient having a score of 2/4, six having scores of 3/4 and the remaining 15 having scores of 4/4. In all cases, these reduced scores were due to artifacts related to rectal gas (Figure 4). This reduced DWI quality compared to 3T resulted in lower PI-QUAL scores (p = 0.0065). Fourteen participants had an overall PI-QUAL score of 3/3, seven participants had a score of 2/3, and one had a score of 1/3. Thus, out of 22 patients, 1 (5%) resulted in a non-diagnostic scan which would need to be repeated.

**Figure 4:**
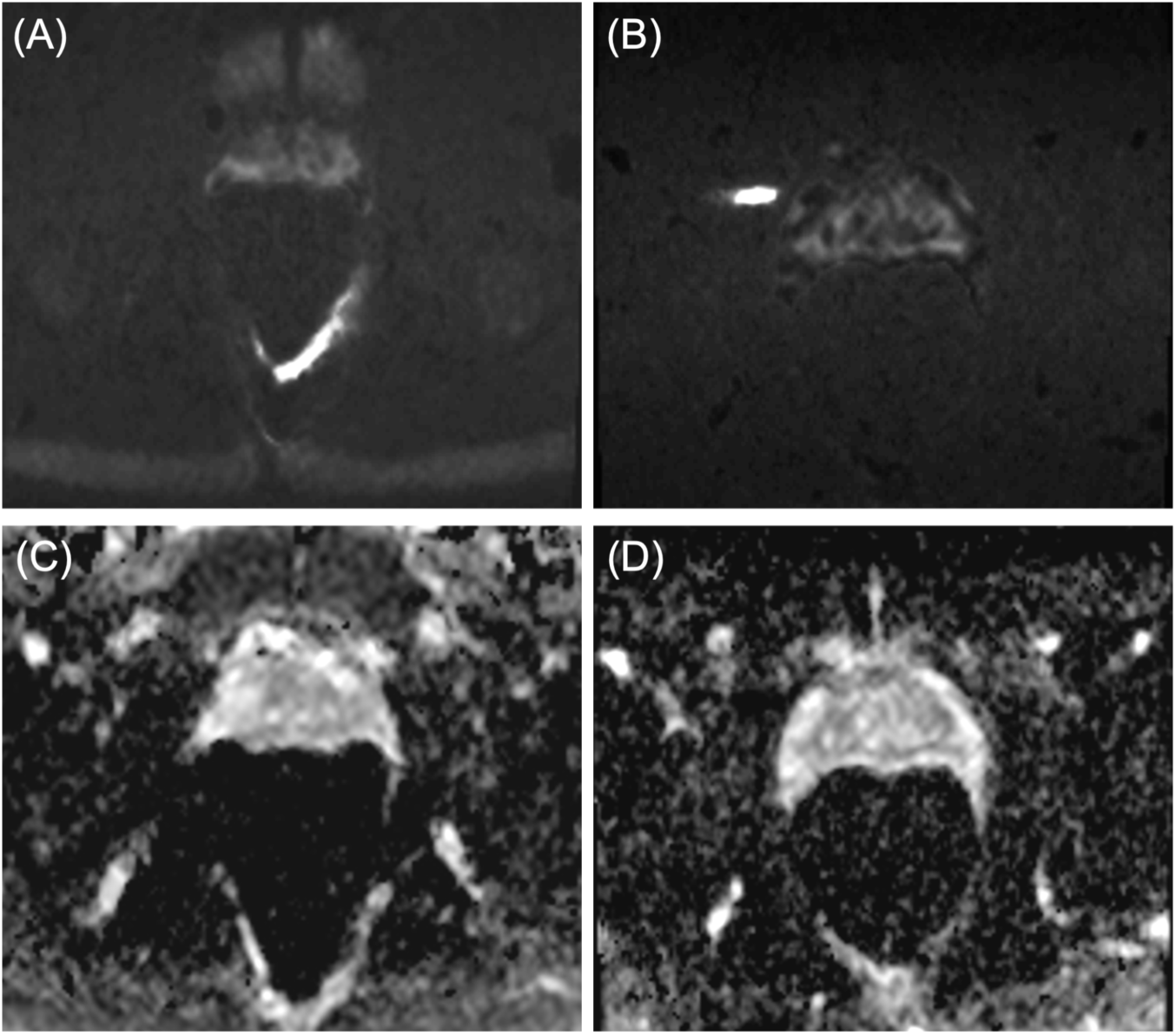
Examples of a volunteer (A and C) and a patient (B and D) with reduced image quality on DWI due to rectal gas on the calculated b-value image at b = 1500 s mm^-2^ (A and B) and ADC map (C and D).

**Table 3:** Image quality scores in patients at 3T and low-field. Bonferroni-corrected p-values are shown from two-tailed Mann-Whitney U-tests.

|  | Score | 3T: n (%) | 0.55T: n (%) | p-value |
| --- | --- | --- | --- | --- |
| T2-WI | 0 | 0 (0) | 0 (0) | 0.4859 |
|  | 1 | 0 (0) | 0 (0) |  |
|  | 2 | 0 (0) | 0 (0) |  |
|  | 3 | 0 (0) | 2 (9) |  |
|  | 4 | 22 (100) | 20 (91) |  |
| DWI | 0 | 0 (0) | 0 (0) | 0.0140 |
|  | 1 | 0 (0) | 0 (0) |  |
|  | 2 | 0 (0) | 1 (5) |  |
|  | 3 | 0 (0) | 6 (27) |  |
|  | 4 | 22 (100) | 15 (68) |  |
| PI-QUAL | 1 | 0 (0) | 1 (5) | 0.0065 |
|  | 2 | 0 (0) | 7 (32) |  |
|  | 3 | 22 (100) | 14 (64) |  |

There was almost perfect agreement between raters for the PI-QUAL score assessment in the control cohort (Cohen’s kappa = 0.96, 95% CI [0.88, 1.0], n = 51) and strong agreement in the patient cohort (Cohen’s kappa = 0.81, 95% CI [0.64, 0.98], n = 44).

### Lesion Detection

Across the 22 patients with suspected PCa, there were 33 lesions detected at 3T. Figure 5 shows a representative example of a lesion seen by both readers at low-field, in comparison with 3T. Figure 6 shows an example of a lesion which was detected at 3T but not seen by either reader at low-field. At low-field, rater 1 identified 26 of these and did not label any false positives, giving a sensitivity of 79%. Rater 2 identified 24 of these and labelled three false positives, giving a sensitivity of 75%. At the patient level (identifying patients with at least one lesion), both raters correctly identified 20 patients and 15 controls, but had two false negatives, giving a sensitivity of 91% and specificity of 100%. Among false negatives, one was overlapping between raters, resulting in strong agreement between raters (Cohen’s kappa = 0.89, 95% CI [0.74, 1.0], n = 37). For both raters, there was no significant difference in PCa detection between low-field and 3T (p = 0.5).

**Figure 5:**
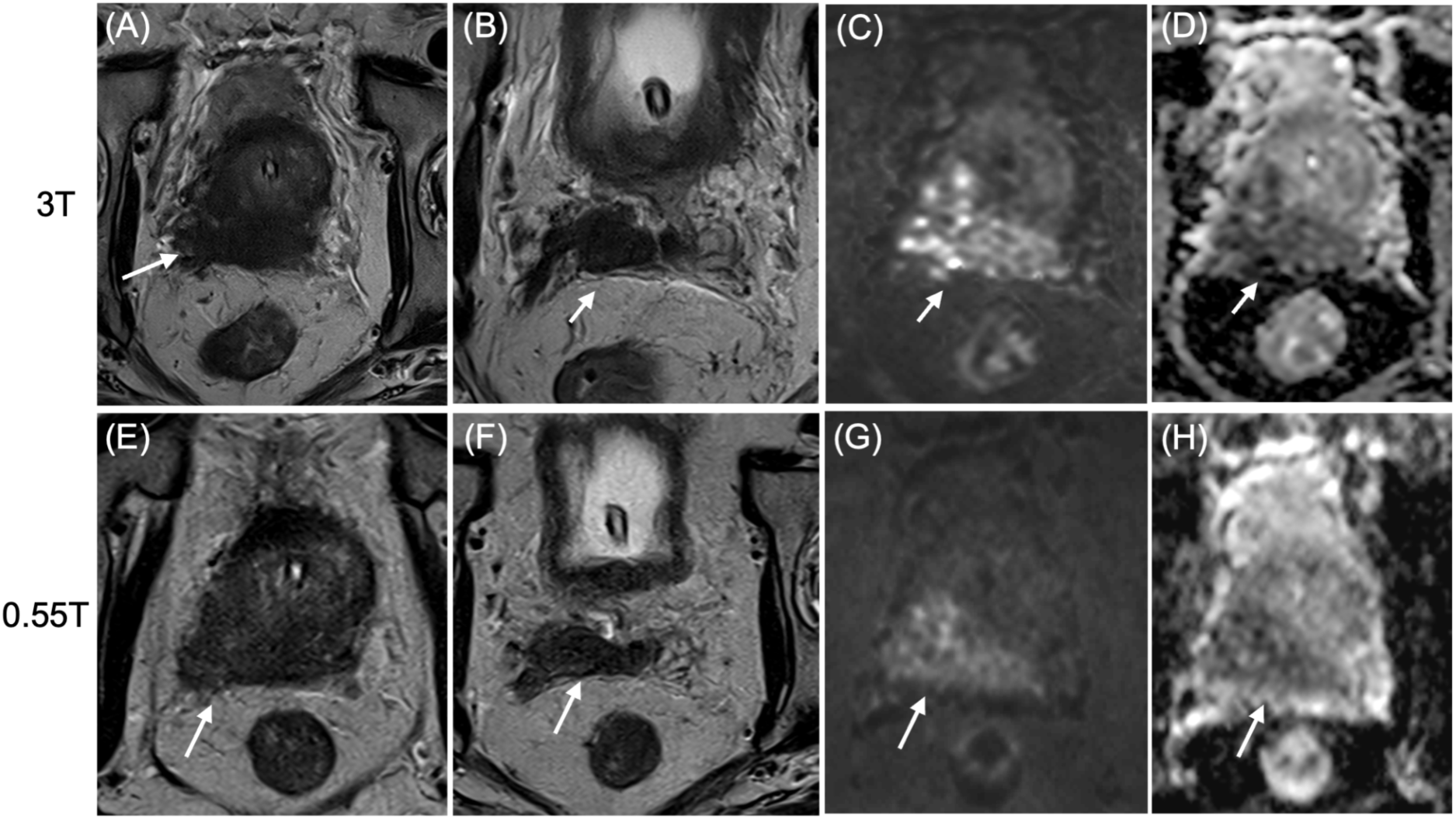
Patient with PI-RADS 5 lesion on 3T prostate MRI (first row, A-D) and corresponding images on 0.55T MRI (second row, E-H). A large lesion developed in the peripheral zone on the right base of the prostate is visible on T2-WI (A and E, arrow), calculated b-value image at b = 1500 s mm^-2^ (C and G, arrow) and ADC map (D and H) with extension to the right seminal vesicle shown on T2-WI (B & F, arrow). The lesion was seen by both readers on low field.

**Figure 6:**
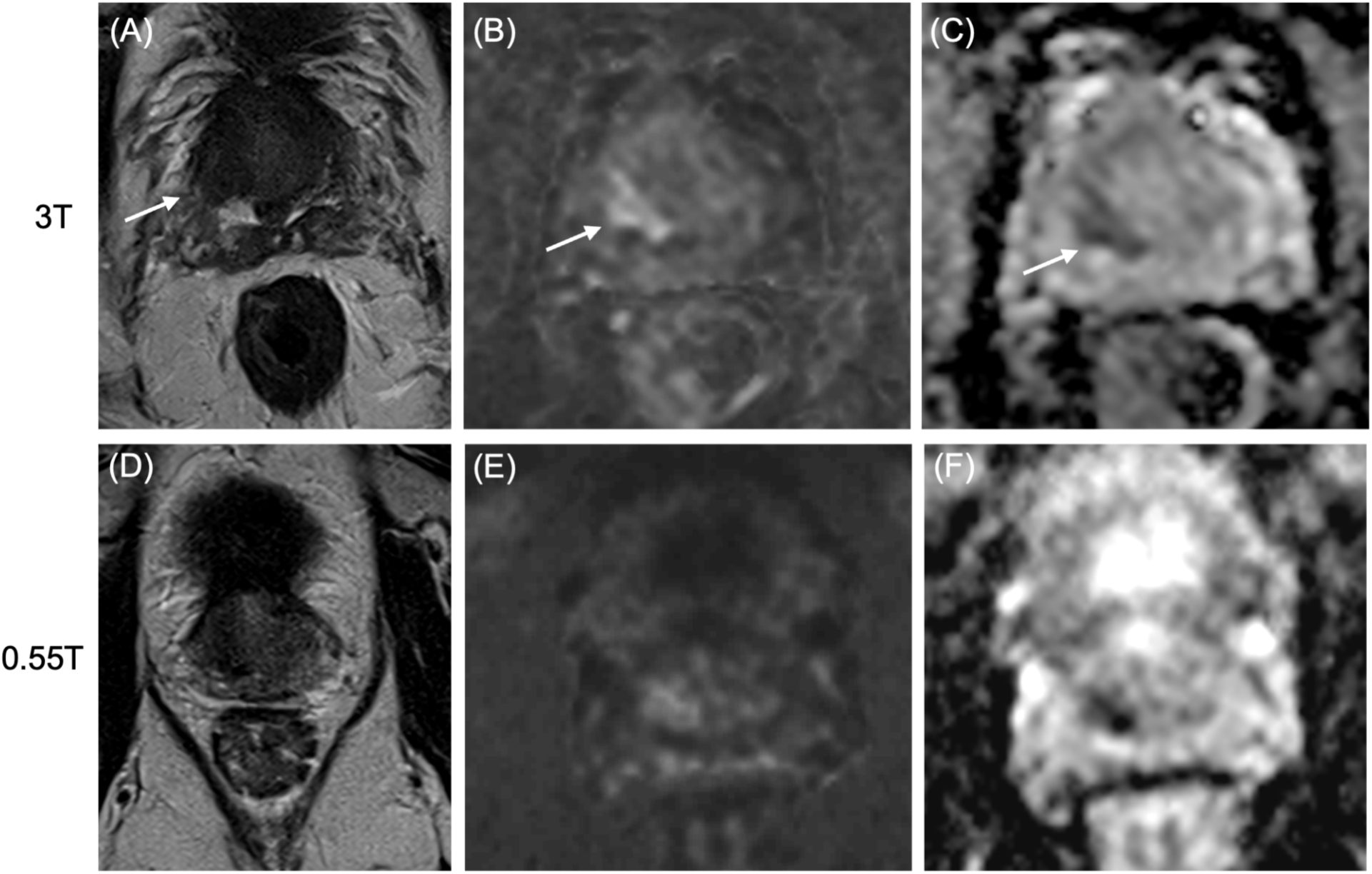
Patient with two PI-RADS 4 lesions on 3T prostate MRI (first row, A–C) and corresponding images on 0.55T MRI (second row, D–F). A right transitional zone lesion was seen on 3T on T2-WI (A, arrow), high b-value (B, arrow) and ADC map (C, arrow) but was not seen by any reader on 0.55T (D–F). Note that the patient had a second lesion that was seen by both readers on 0.55T (not shown here).

## Discussion

The present study demonstrates the feasibility of prostate MRI at low-field. We proposed a clinical prostate MRI protocol at 0.55T, which adheres to the PI-RADS version 2.1 guidelines, exhibits image quality metrics comparable to higher field MRI [14–17, 24–29], and provides promising lesion detection performance compared to 3T.

At 4:02 (minutes:seconds) for axial T2-WI and 6:32 for DWI, the acquisition time of our protocol is substantially shorter than that reported in a comparative study at low-field, which described a duration of 5:11 for T2-WI and 9:28 for DWI with b-values of 50 and 800 [9]. The acquisition time of our T2-WI protocol is comparable to the scan durations mentioned in a review following PI-RADS v2.1 guidelines, of 3:34 and 4:21 at 1.5T and 3T, respectively [30]. The short T2-WI duration was enabled by DLR, as otherwise more averaging would have been needed to achieve sufficient image quality. However, our DLR-enhanced DWI scan was longer than reported in the same review (4:28 and 5:41 minutes at 1.5T and 3T, respectively) [30]. This could be due to the weaker gradient system on the 0.55T which translates into longer diffusion encoding durations and echo planar imaging (EPI) read-out durations. Given the low SNR at 0.55T, image acceleration was used parsimoniously. Our DWI scan time is however still largely within clinically acceptable durations, particularly as it is combined with a shorter T2-WI sequence.

A deep learning denoising and reconstruction method was employed for both T2-WI and DWI data. SNR and CNR were substantially higher in T2-WI with DLR compared to without, as well as higher PI-QUAL scores based on qualitative assessment. A formal comparison between DWI with and without DLR was not made due to unavailable raw DWI for retrospective reconstruction. However, raw DWI scans performed during initial piloting for this project yielded poor image quality with the current acquisition settings, which prompted us to include DLR systematically. These considerations, combined with our observations on the importance of developing bi-parametric protocols with comparable scan times to higher fields, suggest that the use of DLR is an integral part of the large-scale deployment of low-field prostate MRI exams.

This is consistent with other studies of image quality at 0.55T compared to higher fields, which also promoted DLR for spine or knee imaging [31, 32].

ADC values calculated from b-values of 50 and 800 s mm^-2^ were comparable to those calculated from b-values of 50 and 1000 s mm^-2^, suggesting the higher SNR at b = 800 s mm^-2^ can be leveraged to improve image quality for the visual assessment of DWI contrast by the radiologist, without a loss of precision in ADC. Qualitative assessment also suggested a trend towards improved image quality with 800 s mm^-2^ compared to 1000 s mm^-2^. For DWI with b-values of 50 and 800 s mm^-2^, the calculated b-value of 1500 s mm^-^² showed the highest SNR among the tested calculated b-values. A previous study conducted on low-field MRI also demonstrated that the calculated b-value of 1500 s mm^-^² enabled the detection of tumour lesions in two patients [33], further supporting this as a suitable choice for future prostate cancer screening protocols at 0.55T.

SNR of T2-WI (ranging 4–18 in our study) was comparable to values previously reported from sequences at 3T and 1.5T (reported values range from 4–34) [14, 16, 17, 24–28]. Even without DLR, CNR between the PZ and TZ was comparable to previously reported values at 3T (reported median values range from 2-3, similarly to our data) [14, 24]. This CNR was greatly enhanced by DLR (median value of 5.84). In DWI images, SNR was comparable to values at 3T for trace-weighted images at similar b-values; Lee et al., reported SNR values in the range 22– 34 for b = 100 s mm^-2^ and 15–27 for b = 750 s mm^-2^ [14]. Note that these are apparent SNR measurements which do not account for spatially varying noise due to parallel imaging acceleration, therefore values from previous studies provide a contextual reference rather than strict comparison. Median ADC values in the PZ and TZ, of 1.70 and 1.44 x10^-3^ mm^2^ s^-1^ respectively, were higher than those reported by Segeroth et al. at 0.55T (1.50 and 1.34 x10^-3^ mm^2^ s^-1^ in the PZ and central gland, respectively) [9], but were within previously reported ranges at 3T (1.1–1.7 x10^-3^ mm^2^ s^-1^) [14, 15, 29], and offered good contrast between the PZ and TZ. Further study is needed to determine whether this translates to good contrast between malignant and benign tissue.

Among the subjects who received a PI-QUAL score less than 3/3, the limitation was primarily attributable to low DWI scores, mainly due to artifacts caused by rectal gas. In one patient, gas artifacts even resulted in a PI-QUAL score of 1/3, representing a 5% rate of insufficient quality scans in patients (or 3% of the entire cohort). This compares favourably with previous work at 3T, in which 4% of examinations were considered insufficient for diagnostic purposes [34]. To note, all comparative 3T scans were performed with prior enema as it is part of our clinical routine. We suggest that enema should, for now, be recommended also before 0.55T prostate MRI, to minimise artifacts, particularly in DWI, which may further enhance the overall PI-QUAL score [35, 36]. In future, the development of alternative image readouts may not only provide diagnostic image quality without prior enema, but also in patients with metallic implants. For example, segmented EPI in the read-out direction [37] or spiral read-outs are advantageous at low-field and have already been explored for other contrasts and applications [38–40]. Stronger gradients with higher slew rate (more comparable to 3T systems) will also contribute to reducing spatial distortions.

Our data on lesion detection suggest that low-field MRI can identify most lesions visible at 3T, although lesion-level sensitivity was lower than the patient-level sensitivity. The two readers identified 79% and 73% of the 33 lesions detected at 3T. At the patient level, there was a high sensitivity of 91%, and the high inter-reader agreement supports the reproducibility of lesion detection at low-field. These findings should be interpreted in the context of the reference used in this study; the reported performance represents agreement with lesion detection at 3T rather than sensitivity for PCa. Similarly, the lesions which were identified at 3T but not seen at low-field (and therefore counted as false negatives) may in fact be true negatives. Importantly, there was no significant difference between the low-field and the 3T evaluation. Recent meta-analyses reported that patient-level sensitivity and specificity for PI-RADS ≥4 was 88% and 66%, respectively [41, 42]. Thus, the high correspondence between patient-level evaluations at low-field and 3T may translate to a high sensitivity for PCa detection, but further work with biopsy as the gold standard is required to establish true clinically significant PCa detection performance. Taken together, these initial findings indicate that, despite some reduction in lesion-level detection compared with 3T, the optimised low-field protocol generally provides diagnostically usable examinations and preserves patient-level detection of suspicious lesions.

### Limitations

As the aims of this study were to optimise a low-field prostate MRI protocol and demonstrate the feasibility of lesion detection at low-field, the sample size was small. A larger sample size may provide more diversity in terms of metallic implants, enabling better characterisation of susceptibility distortions.

Additionally, the SNR calculation we used does not account for spatially varying noise statistics due to parallel imaging and multichannel coil reconstruction [43–45]. Consequently, this does not represent the true physical SNR [46, 47], but rather provides a measure of apparent SNR for the purposes of evaluating the effective signal-to-noise characteristics of the reconstructed image as presented to the reader. While this approach is appropriate for relative comparisons between sequences within the context of this study, direct quantitative comparison with previous studies is limited by differences in acquisition parameters and reconstruction techniques. As such, previously reported SNR ranges derived using equivalent ROI-based methods are presented for contextual reference rather than strict comparison.

Due to technical constraints, retrospective reconstructions without DLR were not available for DWI data. This unfortunately prevented a direct comparison of SNR in DWI with and without DLR.

Finally, we did not have biopsy data available as a ground truth for lesion detection analysis. Therefore, we could not quantify the rate of PCa detection for each field strength. Instead, we opted to use 3T as a reference to determine whether low-field lesion detection was comparable to a standard clinical evaluation.

## Conclusion

We proposed a bi-parametric prostate MRI protocol at 0.55T, adhering to PI-RADS technical recommendations. This protocol had similar scan time compared to its 3T and 1.5T counterparts, while DLR ensured high image quality which provided a promising lesion detection rate compared to 3T. Thus, low-field MRI shows promise for PCa screening, warranting further validation of PI-RADS performance against pathology-confirmed lesions and in a PCa screening population.

## Data Availability

Data are available upon reasonable request to the corresponding author.

## Acknowledgments

The authors thank Omar Darwish and Marco Mueller (Siemens Healthineers) for assistance with the MRI sequences. This study received funding from a Seed Grant of the Department of Radiology, Lausanne University Hospital, as well as the Swiss Secretariat for Research and Innovation (SERI) Starting Grant award ‘FIREPATH’ MB22.00032, Swiss National Science Foundation #194260.

## Data Availability Statement

The data presented in this work are available upon reasonable request.

## References

1. Yilmaz EC, Esengur OT, Gelikman DG, Turkbey B (2025) Interpreting Prostate Multiparametric MRI: Beyond Adenocarcinoma – Anatomical Variations, Mimickers, and Post-Intervention Changes. Semin Ultrasound CT MRI 46:2–30. 10.1053/j.sult.2024.11.001

2. Williams C, Khondakar N, Pinto P, Turkbey B (2021) The Importance of Quality in Prostate MRI. Semin Roentgenol 56:384–390. 10.1053/j.ro.2021.08.005

3. Weinreb JC, Barentsz JO, Choyke PL, et al (2016) PI-RADS Prostate Imaging – Reporting and Data System: 2015, Version 2. Eur Urol 69:16–40. 10.1016/j.eururo.2015.08.052

4. Chandarana H, Bagga B, Huang C, et al (2021) Diagnostic abdominal MR imaging on a prototype low-field 0.55 T scanner operating at two different gradient strengths. Abdom Radiol 46:5772–5780. 10.1007/s00261-021-03234-1

5. Shetty AS, Ludwig DR, Ippolito JE, et al (2023) Low-Field-Strength Body MRI: Challenges and Opportunities at 0.55 T. RadioGraphics 43:e230073. 10.1148/rg.230073

6. Giganti F, Allen C, Emberton M, et al (2020) Prostate Imaging Quality (PI-QUAL): A New Quality Control Scoring System for Multiparametric Magnetic Resonance Imaging of the Prostate from the PRECISION trial. Eur Urol Oncol 3:615–619. 10.1016/j.euo.2020.06.007

7. De Rooij M, Allen C, Twilt JJ, et al (2024) PI-QUAL version 2: an update of a standardised scoring system for the assessment of image quality of prostate MRI. Eur Radiol 34:7068– 7079. 10.1007/s00330-024-10795-4

8. Makwana RD, Wunsch DC, Punwani S, et al (2026) Current state of the art of new prostate MRI technologies and potential future developments. BJR|Open 8:tzag011. 10.1093/bjro/tzag011

9. Segeroth M, Breit H-C, Wasserthal J, et al (2025) AI-Based Evaluation of Prostate MR Imaging at a Modern Low-field 0.55 T Scanner Compared to 3 T in a Screening Cohort. Acad Radiol 32:2700–2706. 10.1016/j.acra.2024.11.024

10. Kaur T, Jiang Y, Seiberlich N, et al (2025) Clinical feasibility of MRI-guided in-bore prostate biopsies at 0.55T. Abdom Radiol 50:3773–3783. 10.1007/s00261-024-04783-x

11. Seifert AC, Obmann MM, Breit H-C, et al (2025) Deep Learning-Enhanced Diffusion-Weighted Imaging of the Abdomen at 0.55 T: Image Quality and Apparent Diffusion Coefficient Calculation Interchangeability in Healthy Volunteers. Acad Radiol S1076633225011250. 10.1016/j.acra.2025.11.039

12. Ramachandran A, Hussain HK, Gulani V, et al (2024) Abdominal MRI on a Commercial 0.55T System: Initial Evaluation and Comparison to Higher Field Strengths. Acad Radiol 31:3177–3190. 10.1016/j.acra.2024.01.018

13. Koonjoo N, Zhu B, Bagnall GC, et al (2021) Boosting the signal-to-noise of low-field MRI with deep learning image reconstruction. Sci Rep 11:8248. 10.1038/s41598-021-87482-7

14. Lee K-L, Kessler DA, Dezonie S, et al (2023) Assessment of deep learning-based reconstruction on T2-weighted and diffusion-weighted prostate MRI image quality. Eur J Radiol 166:111017. 10.1016/j.ejrad.2023.111017

15. Ueda T, Ohno Y, Yamamoto K, et al (2022) Deep Learning Reconstruction of Diffusion-weighted MRI Improves Image Quality for Prostatic Imaging. Radiology 303:373–381. 10.1148/radiol.204097

16. Harder FN, Weiss K, Amiel T, et al (2022) Prospectively Accelerated T2-Weighted Imaging of the Prostate by Combining Compressed SENSE and Deep Learning in Patients with Histologically Proven Prostate Cancer. Cancers 14:5741. 10.3390/cancers14235741

17. Park JC, Park KJ, Park MY, et al (2022) Fast T2-Weighted Imaging With Deep Learning-Based Reconstruction: Evaluation of Image Quality and Diagnostic Performance in Patients Undergoing Radical Prostatectomy. J Magn Reson Imaging 55:1735–1744. 10.1002/jmri.27992

18. Bischoff LM, Peeters JM, Weinhold L, et al (2023) Deep Learning Super-Resolution Reconstruction for Fast and Motion-Robust T2-weighted Prostate MRI. Radiology 308:e230427. 10.1148/radiol.230427

19. Turkbey B, Rosenkrantz AB, Haider MA, et al (2019) Prostate Imaging Reporting and Data System Version 2.1: 2019 Update of Prostate Imaging Reporting and Data System Version 2. Eur Urol 76:340–351. 10.1016/j.eururo.2019.02.033

20. Liu W, Darwish O, Benkert T, et al (2024) Improved readout-segmented EPI using deep learning reconstruction. In: Proceedings of the International Society for Magnetic Resonance in Medicine. Singapore, p 5106

21. Klein S, Staring M, Murphy K, et al (2010) elastix: A Toolbox for Intensity-Based Medical Image Registration. IEEE Trans Med Imaging 29:196–205. 10.1109/TMI.2009.2035616

22. Shamonin D (2013) Fast parallel image registration on CPU and GPU for diagnostic classification of Alzheimer’s disease. Front Neuroinformatics 7:. 10.3389/fninf.2013.00050

23. Li X, Huang W, Rooney WD (2012) Signal-to-noise ratio, contrast-to-noise ratio and pharmacokinetic modeling considerations in dynamic contrast-enhanced magnetic resonance imaging. Magn Reson Imaging 30:1313–1322. 10.1016/j.mri.2012.05.005

24. Ullrich T, Quentin M, Oelers C, et al (2017) Magnetic resonance imaging of the prostate at 1.5 versus 3.0 T: A prospective comparison study of image quality. Eur J Radiol 90:192– 197. 10.1016/j.ejrad.2017.02.044

25. Lewis S, Ganti A, Argiriadi P, et al (2022) Prostate MRI using a rigid two-channel phased-array endorectal coil: comparison with phased array coil acquisition at 3 T. Cancer Imaging 22:15. 10.1186/s40644-022-00453-7

26. O’Donohoe RL, Dunne RM, Kimbrell V, Tempany CM (2019) Prostate MRI using an external phased array wearable pelvic coil at 3T: comparison with an endorectal coil. Abdom Radiol 44:1062–1069. 10.1007/s00261-018-1804-9

27. Barth BK, Cornelius A, Nanz D, et al (2016) Comparison of image quality and patient discomfort in prostate MRI: pelvic phased array coil vs. endorectal coil. Abdom Radiol 41:2218–2226. 10.1007/s00261-016-0819-3

28. Kim BS, Kim T-H, Kwon TG, Yoo ES (2012) Comparison of Pelvic Phased-Array versus Endorectal Coil Magnetic Resonance Imaging at 3 Tesla for Local Staging of Prostate Cancer. Yonsei Med J 53:550. 10.3349/ymj.2012.53.3.550

29. Mazaheri Y, Vargas HA, Nyman G, et al (2013) Diffusion-weighted MRI of the prostate at 3.0T: Comparison of endorectal coil (ERC) MRI and phased-array coil (PAC) MRI—The impact of SNR on ADC measurement. Eur J Radiol 82:e515–e520. 10.1016/j.ejrad.2013.04.041

30. Purysko AS, Baroni RH, Giganti F, et al (2021) PI-RADS Version 2.1: A Critical Review, From the *AJR* Special Series on Radiology Reporting and Data Systems. Am J Roentgenol 216:20–32. 10.2214/AJR.20.24495

31. Lopez Schmidt I, Haag N, Shahzadi I, et al (2023) Diagnostic Image Quality of a Low-Field (0.55T) Knee MRI Protocol Using Deep Learning Image Reconstruction Compared with a Standard (1.5T) Knee MRI Protocol. J Clin Med 12:1916. 10.3390/jcm12051916

32. Schlicht F, Vosshenrich J, Donners R, et al (2024) Advanced deep learning-based image reconstruction in lumbar spine MRI at 0.55 T – Effects on image quality and acquisition time in comparison to conventional deep learning-based reconstruction. Eur J Radiol Open 12:100567. 10.1016/j.ejro.2024.100567

33. Lemberskiy G, Chandarana H, Bruno M, et al (2023) Feasibility of Accelerated Prostate Diffusion-Weighted Imaging on 0.55 T MRI Enabled With Random Matrix Theory Denoising. Invest Radiol 58:720–729. 10.1097/RLI.0000000000000979

34. Karanasios E, Caglic I, Zawaideh JP, Barrett T (2022) Prostate MRI quality: clinical impact of the PI-QUAL score in prostate cancer diagnostic work-up. Br J Radiol 95:20211372. 10.1259/bjr.20211372

35. Schmidt C, Hötker AM, Muehlematter UJ, et al (2021) Value of bowel preparation techniques for prostate MRI: a preliminary study. Abdom Radiol 46:4002–4013. 10.1007/s00261-021-03046-3

36. Lin Y, Yilmaz EC, Belue MJ, Turkbey B (2023) Prostate MRI and image Quality: It is time to take stock. Eur J Radiol 161:110757. 10.1016/j.ejrad.2023.110757

37. Liney GP, Holloway L, Al Harthi TM, et al (2015) Quantitative evaluation of diffusion-weighted imaging techniques for the purposes of radiotherapy planning in the prostate. Br J Radiol 88:20150034. 10.1259/bjr.20150034

38. Bauman G, Lee NG, Tian Y, et al (2023) Submillimeter lung MRI at 0.55 T using balanced steady-state free precession with half-radial dual-echo readout (bSTAR). Magn Reson Med 90:1949–1957. 10.1002/mrm.29757

39. Tian Y, Nayak KS (2024) Real-time water/fat imaging at 0.55T with spiral out-in-out-in sampling. Magn Reson Med 91:649–659. 10.1002/mrm.29885

40. Ramasawmy R, Herzka DA, Restivo MC, et al (2019) Spin-echo imaging at 0.55T using a spiral trajectory. In: Proceedings of the International Society for Magnetic Resonance in Medicine. Montreal, p 1192

41. Oerther B, Nedelcu A, Engel H, et al (2024) Update on PI-RADS Version 2.1 Diagnostic Performance Benchmarks for Prostate MRI: Systematic Review and Meta-Analysis. Radiology 312:e233337. 10.1148/radiol.233337

42. Nedelcu A, Oerther B, Benkendorff A, et al (2026) PI-RADS Version 2.1 for Prostate MRI Interpretation: Associations of Study Quality and Cancer Detection Metrics—A Systematic Review and Meta-Analysis. Am J Roentgenol 226:e2533583. 10.2214/AJR.25.33583

43. Deshmane A, Gulani V, Griswold MA, Seiberlich N (2012) Parallel MR imaging. J Magn Reson Imaging 36:55–72. 10.1002/jmri.23639

44. Dietrich O, Raya JG, Reeder SB, et al (2007) Measurement of signal-to-noise ratios in MR images: Influence of multichannel coils, parallel imaging, and reconstruction filters. J Magn Reson Imaging 26:375–385. 10.1002/jmri.20969

45. Aja-Fernández S, Vegas-Sánchez-Ferrero G, Tristán-Vega A (2014) Noise estimation in parallel MRI: GRAPPA and SENSE. Magn Reson Imaging 32:281–290. 10.1016/j.mri.2013.12.001

46. Reeder SB, Wintersperger BJ, Dietrich O, et al (2005) Practical approaches to the evaluation of signal-to-noise ratio performance with parallel imaging: Application with cardiac imaging and a 32-channel cardiac coil. Magn Reson Med 54:748–754. 10.1002/mrm.20636

47. Robson PM, Grant AK, Madhuranthakam AJ, et al (2008) Comprehensive quantification of signal-to-noise ratio and *g* -factor for image-based and *k* -space-based parallel imaging reconstructions. Magn Reson Med 60:895–907. 10.1002/mrm.21728

