## Supplementary Materials for "Feasibility of PI-RADS Compliant Prostate MRI at 0.55T"

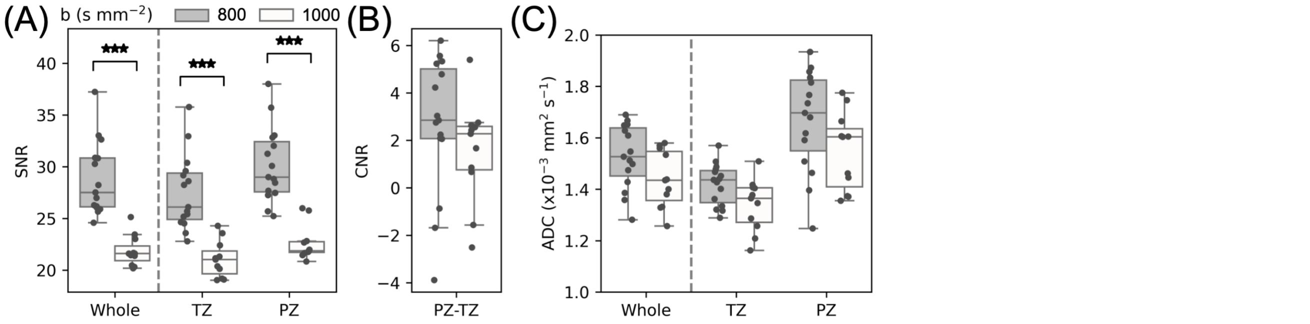


Supplementary Figure 1: Comparison of DWI data acquired with b-values of 50 and 800 s mm$^{-2}$ to data acquired with b-values of 50 and 1000 s mm$^{-2}$. SNR (A) and CNR (B) values are shown for the trace-weighted image from the high b-value in each acquisition. ADC values are shown for each acquisition (C). The data shown here is reproduced from Figure 3 and Supplementary Figure 2 to provide a direct comparison between the two DWI acquisitions. *$p < 0.05$; **$p < 0.01$; ***$p < 0.001$.

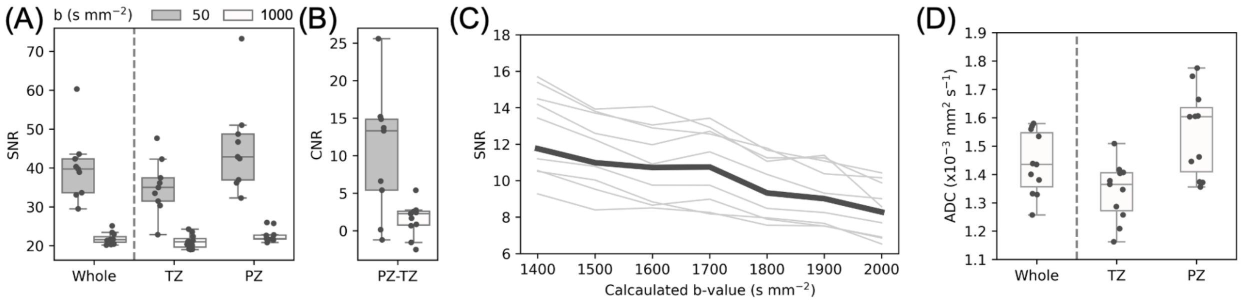


Supplementary Figure 2: Quantitative measurements from DWI data acquired with b-values of 50 and 1000 s mm$^{-2}$ (to be compared with Fig. 3E-H in the main text). SNR (A) and CNR (B) are shown for each acquired b-value for the whole prostate, TZ and PZ. SNR in the whole prostate is shown for calculated b-values in the range 1400–2000 s mm$^{-2}$ (C), showing each subject as a thin grey line, with the median shown as a thick black line. ADC values are shown for the whole prostate, TZ and PZ (D).

Supplementary Table 1: 3T MRI acquisition parameters.

| | **T2-WI** (axial / sagittal / coronal) | **DWI** |
|---|---|---|
| TR (ms) | 7740 / 7440 / 7440 | 4500 |
| TE (ms) | 104 | 65 |
| Resolution (mm) | 0.3 x 0.3 | 0.9 x 0.9 |
| Slice thickness (mm) | 3.0 | 3.0 |
| Matrix | 384 x 384 (acquisition matrix)<br>768 x 768 (reconstruction) | 114 x 58 (acquisition matrix)<br>228 x 116 (reconstruction) |
| Field of view (mm) | 200 x 200 | 200 x 102 |
| Number of slices | 26 / 25 / 25 | 25 |
| Acceleration | GRAPPA 4 | Partial Fourier 7/8 |
| Turbo factor | 25 | n/a |
| b-values (s $mm^{-2}$) | n/a | 50, 1000 |
| Number of signal averages | 1 | 2, 10 |
| Acquisition time (minutes : seconds) | 1:46 / 1:34 / 1:42 | 3:47 |

Supplementary Table 2: Qualitative image scores in healthy controls from each rater. DWI scores were assigned for each sequence based on the ADC map and the calculated b-value image with the highest SNR (b = 1500 s $mm^{-2}$ for $DWI_{800}$ and b = 1400 s $mm^{-2}$ for $DWI_{1000}$).

| | Score | Rater 1: n (%) | Rater 2: n (%) |
|---|---|---|---|
| $DWI_{800}$ | 0 | 0 (0) | 0 (0) |
| | 1 | 0 (0) | 0 (0) |
| | 2 | 0 (0) | 0 (0) |
| | 3 | 5 (33) | 5 (33) |
| | 4 | 10 (67) | 10 (67) |
| $DWI_{1000}$ | 0 | 0 (0) | 0 (0) |
| | 1 | 0 (0) | 0 (0) |
| | 2 | 0 (0) | 0 (0) |
| | 3 | 8 (73) | 6 (55) |
| | 4 | 3 (28) | 5 (45) |
| T2-WI raw | 0 | 0 (0) | 0 (0) |
| | 1 | 0 (0) | 0 (0) |
| | 2 | 0 (0) | 0 (0) |
| | 3 | 10 (100) | 10 (100) |
| | 4 | 0 (0) | 0 (0) |
| T2-WI DLR | 0 | 0 (0) | 0 (0) |
| | 1 | 0 (0) | 0 (0) |
| | 2 | 0 (0) | 0 (0) |
| | 3 | 1 (7) | 1 (7) |
| | 4 | 14 (93) | 14 (93) |

Supplementary Table 3: Consensus image quality scores at two b-values for DWI, and with and without DLR for T2-WI, shown as median (IQR). DWI scores were assigned for each sequence based on the ADC map and the calculated b-value image with the highest SNR (b = 1500 s $mm^{-2}$ for $DWI_{800}$ and b = 1400 s $mm^{-2}$ for $DWI_{1000}$). [a]Two-tailed Mann-Whitney U-test. [b]One-tailed Wilcoxon signed-rank test.

| | Score | b = [50, 800] (s $mm^2$): n (%) | b = [50, 1000] (s $mm^2$): n (%) | p-value |
|---|---|---|---|---|
| DWI score | 0 | 0 (0) | 0 (0) | 0.055[a] |
| | 1 | 0 (0) | 0 (0) | |
| | 2 | 0 (0) | 0 (0) | |
| | 3 | 5 (33) | 8 (73) | |
| | 4 | 10 (67) | 3 (27) | |
| | | non-DLR | DLR | p-value |
| T2-WI score | 0 | 0 (0) | 0 (0) | 0.002[b] |
| | 1 | 0 (0) | 0 (0) | |
| | 2 | 0 (0) | 0 (0) | |
| | 3 | 11 (92) | 1 (7) | |
| | 4 | 1 (8) | 14 (93) | |

Supplementary Table 4: Qualitative image scores in suspected PCa patients from each rater. DWI scores were assigned based on the ADC map and the calculated b-value image at b = 1500 s $mm^{-2}$.

| | Score | Rater 1: n (%) | Rater 2: n (%) |
|---|---|---|---|
| DWI | 0 | 0 (0) | 0 (0) |
| | 1 | 0 (0) | 0 (0) |
| | 2 | 1 (5) | 0 (0) |
| | 3 | 5 (23) | 6 (27) |
| | 4 | 16 (73) | 16 (73) |
| T2-WI | 0 | 0 (0) | 0 (0) |
| | 1 | 0 (0) | 0 (0) |
| | 2 | 0 (0) | 0 (0) |
| | 3 | 2 (9) | 2 (9) |
| | 4 | 20 (91) | 20 (91) |
